# Motor phenotypes of amyotrophic lateral sclerosis – a three-determinant anatomical classification based on the region of onset, propagation of motor symptoms, and the degree of upper and lower motor neuron dysfunction

**DOI:** 10.1101/2025.03.21.25324256

**Authors:** Thomas Meyer, Matthias Boentert, Julian Großkreutz, Patrick Weydt, Sarah Bernsen, Peter Reilich, Robert Steinbach, Annekathrin Rödiger, Joachim Wolf, Ute Weyen, Albert C. Ludolph, Jochen Weishaupt, Susanne Petri, Paul Lingor, René Günther, Wolfgang Löscher, Markus Weber, Christoph Münch, André Maier, Torsten Grehl

## Abstract

**Background:** In amyotrophic lateral sclerosis (ALS), heterogeneity of motor phenotypes is a fundamental hallmark of the disease. Distinct ALS phenotypes were associated with a different progression and survival. Despite its relevance for clinical practice and research, there is no broader consensus on the classification of ALS phenotypes.

**Methods:** An expert consensus process for the classification of ALS motor phenotypes was performed from May 2023 to December 2024. A three-determinant anatomical classification was proposed which is based on the 1) region of onset (O), 2) the propagation of motor symptoms (P), and 3) the degree of upper (UMN) and/or lower motor neuron (LMN) dysfunction (M). Accordingly, this classification is referred to as the “OPM classification”.

**Results:** Onset phenotypes differentiate the site of first motor symptoms: O1) head onset; O2d) distal arm onset; O2p) proximal arm onset; O3r) trunk respiratory onset; O3a) trunk axial onset; O4d) distal leg onset; O4p) proximal leg onset. Propagation phenotypes differentiate the temporal propagation of motor symptoms from the site of onset to another, vertically distant body region: PE) earlier propagation (within 12 months of symptom onset); PL) later propagation (without propagation within 12 months of symptom onset), including the established phenotypes of “progressive bulbar paralysis” (O1, PL), “flail-arm syndrome” (O2p, PL), and “flail-leg syndrome” (O4d, PL); PN) propagation not yet classifiable as time since symptom onset is less than 12 months. Phenotypes of motor neuron dysfunction differentiate the degree of UMN and/or LMN dysfunction: M0) balanced UMN and LMN dysfunction; M1d) dominant UMN dysfunction; M1p) pure UMN dysfunction (“primary lateral sclerosis”, PLS); M2d) dominant LMN dysfunction; M2p) pure LMN dysfunction (“progressive muscle atrophy”, PMA); M3) dissociated motor neuron dysfunction with dominant LMN and UMN dysfunction of the arms and legs (“brachial amyotrophic spastic paraparesis”), respectively.

**Conclusion:** This consensus process aimed to standardize the clinical description of ALS motor phenotypes in clinical practice and research – based on the onset region, propagation pattern, and motor neuron dysfunction. This “OPM classification” contributes to specifying the prognosis, to defining the inclusion or stratification criteria in clinical trials and to correlate phenotypes with the underlying disease mechanisms of ALS.

## BACKGROUND

In amyotrophic lateral sclerosis (ALS), heterogeneity of motor phenotypes is a fundamental hallmark of the disease and is determined by variability of three anatomical determinants: the region of onset; the spatial and temporal propagation of motor dysfunction from the site of onset to other body regions; and the relative involvement of upper (UMN) and lower (LMN) motor neuron degeneration [1–7]. Distinct ALS phenotypes were associated with different progression and survival [8–15]. Given its predictive capacity, clinical phenotyping is of relevance in both, clinical practice and research [8–17].

The history of ALS can be traced back to the description of clinical phenotypes and their anatomical correlates [18–32]. It included progressive muscle atrophy (PMA), a pure LMN involvement phenotype, and conversely a pure UMN variant, which was named primary lateral sclerosis (PLS) [19–26]. The meaning of distinct propagation patterns was already recognized when progressive bulbar palsy (PBP), the flail-arm syndrome (FAS), the thoracic-onset variant, and the flail-leg syndrome (FLS) were distinguished [27–35]. In principle, the specific motor phenotype is determined by the initial focality at the onset of motor neuron degeneration and the subsequent spread through the anatomy of the UMN and/or LMN [1–3, 36–38]. Clinical assessment remains the gold standard for describing the individual phenotype that evolves over the course of the disease in terms of localization and the highly variable extent of UMN and LMN dysfunction [1–3]. From a clinical perspective, classification of phenotypes can improve the prognostic information and may be associated with specific needs and treatment options [16]. Furthermore, phenotypes have implications for case selection and randomization in clinical trials with the assumption that distinct subcohorts progress differently and respond specifically to treatments [4,6,7].

Despite the relevance of this topic for care and clinical trials, there is no broader consensus on the classification or even naming of ALS phenotypes. In the absence of a clinically useful phenotypic classification system, many neurologists, researchers, and trialists employ informal, non-systematic approaches to the classification of ALS [16, 39–41]. In a previous study, a two-determinant classification of motor ALS phenotypes was introduced, differentiating phenotypes of onset region and the degree of UMN/LMN involvement [42]. This classification was found to correlate with NfL, the ALS progression rate (ALSPR), and survival. Based on the phenotyping experience in this large multicenter study, a revision of the phenotype classification and an even more differentiated three-determinant classification is being proposed. The objective of the revision of the phenotype classification was to (i) collect common and rare ALS motor phenotypes in its various names and wordings, (ii) to categorize existing phenotypes in a three-determinant classification system, (iii) to linguistically adapt and harmonize the newly defined phenotypes, iv) to provide guidelines for motor phenotyping in clinical practice (v) to address the advances and limitations of ALS motor phenotype classification.

## METHODS

### Setting

An expert consensus process for the classification of ALS motor phenotypes was performed from May 2023 to December 2024. The initiative was carried out by a consensus group of the NfL-ALS consortium – a multicenter natural history study aiming at the systematic assessment of clinical characteristics, serum NfL, and ALS motor phenotypes [42, 43]. In this observational study, 16 specialized ALS centers are organized. This consensus group encompassed 20 ALS experts, each with expertise in ALS motor phenotyping.

### Design

A consensus group design was used to evolve consensus for ALS motor phenotypes. The consensus process encompassed the 1) compilation of circulating phenotype classifications including the variable naming and synonymous terms of phenotypes; 2) ordering of existing and newly described phenotypes; 3) iterative discussion of renamed and ordered phenotypes; 4) linguistic harmonization; 5) consensus finding, and 6) release of guidelines for the use of the three-determinant phenotype classification.

### Use cases of phenotyping

The phenotypes are intended to support the following use cases in clinical practice and research: 1) specifying diagnosis in clinical practice; 2) predicting the disease course in patient counseling; 3) defining inclusion, exclusion, and stratification in clinical trials; 4) association with specific genotypes, etiologies, or pathophysiological mechanisms.

### Exclusion of non-motor phenotypes

The classification of phenotypes was limited to those associated with ALS and motor function. Non-motor phenotypes, including cognitive, behavioral, and metabolic symptoms, were not within the scope of this classification nor were they subject to the corresponding consensus process.

## RESULTS

### Assessment of region of onset

Phenotypes of onset differentiate the site of first symptoms including the head, arm, trunk or leg region. First symptoms are defined as impaired motor function including weakness or/and slowed, poorly coordinated voluntary movement, dysarthria, dysphagia, and hypoventilation. Motor neuron signs and symptoms without motor functional impairment (such as fasciculations, muscle wasting, increased or pathological reflexes) are not classified as first symptoms. The assessment is based on the patient history (**Figure 1 and 2**, **Table 1**).

**Figure 1.**
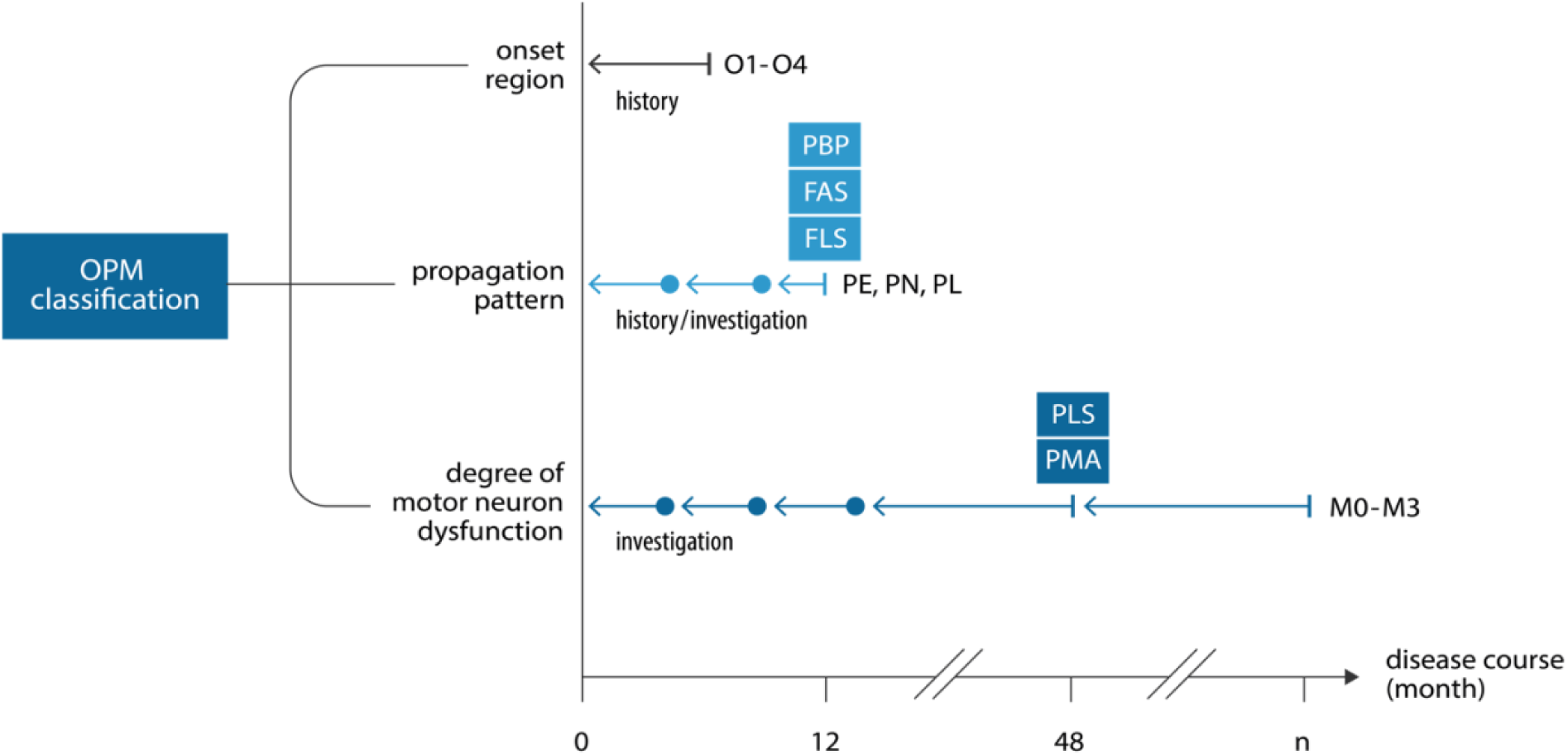
Assessment of ALS motor phenotypes. Phenotypes of the region of onset (O1-4) are assessed by patient history. Phenotypes of propagation (PE, PL, PN) pattern are determined by follow-up of the patient history and/or neurological investigation during the disease course. Some propagation phenotypes can be classified with certainty only after 12 months of follow-up. This includes the phenotypes of late propagation (PL), including the historic descriptions of progressive bulbar palsy (PBP), flail-arm syndrome (FAS) or flail-leg syndrome (FLS), respectively. Phenotypes of the motor neuron dysfunction (M0-M3) are assessed by neurological investigation of the degree of upper (UMN) and/or lower (LMN) neuron involvement. Some motor neuron dysfunction phenotypes can be classified with certainty only after 48 months of follow-up. This includes the phenotypes of pure UMN (M1p) and LMN dysfunction (M2p) in its historic descriptions of primary lateral sclerosis (PLS) and progressive muscle atrophy (PMA). In principle, phenotypes of the motor neuron dysfunction can be changing during the complete disease course. Therefore, motor neuron dysfunction phenotypes need to be re-evaluated as the disease progresses. O1) head onset; O2) arm onset; O3) trunk onset; O4) leg onset; PE) earlier propagation; PL) later propagation; PN) propagation not yet classifiable ; M0) balanced UMN and LMN dysfunction; M1d) dominant UMN dysfunction; M1p) pure UMN dysfunction; M2d) dominant LMN dysfunction; M2p) pure LMN dysfunction; M3) dissociated motor neuron dysfunction with dominant LMN and UMN dysfunction of the arms and legs. Arrow, retrospective assessment period.

**Figure 2.**
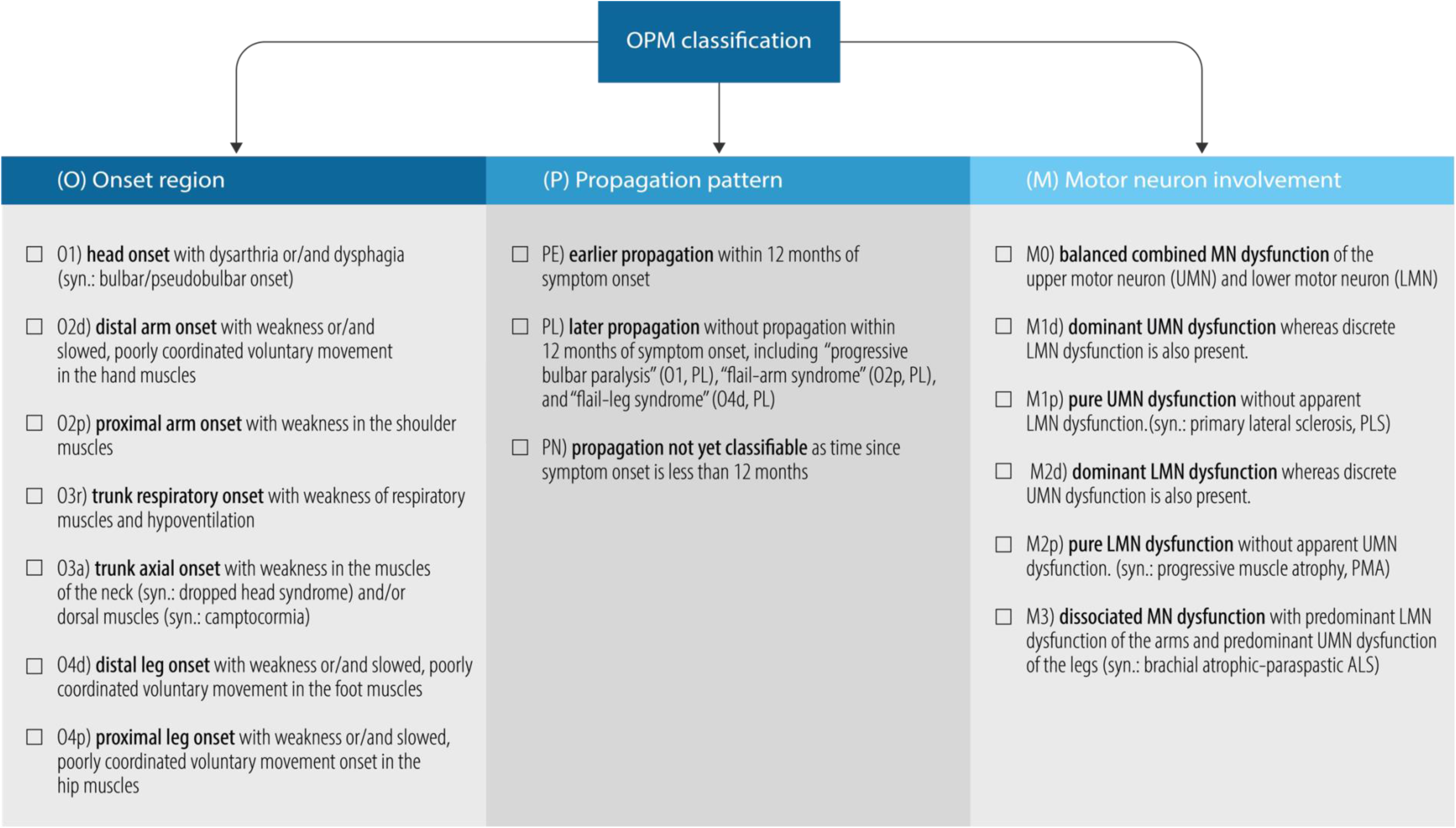
Classification of ALS motor phenotypes. Onset region (O): Phenotypes of onset differentiate the site of first symptoms including the bulbar region, arm, trunk and leg; phenotypes of propagation (P) differentiate the earlier (within 12 months), later (after 12 months) or unclassifiable (monitoring of 12 months not completed) propagation of motor neuron dysfunction from the region of onset to another, vertically distant body region; phenotypes of motor neuron dysfunction (M) differentiate the degree of clinical upper (UMN) and lower motor neuron (LMN) dysfunction.

**Table 1.** ALS motor phenotypes – guidance for assessment.

| ALS motor phenotype | Code | synonymous | Guidance for assessment | Commentary |
| --- | --- | --- | --- | --- |
| **Onset region** |  |  |  |  |
| **head region onset** | O1 | bulbar/pseudobulbar onset | Dysarthria or/and dysphagia are first motor symptoms which reflect weakness or/and slowed, poorly coordinated voluntary movement of the tongue and palatal muscles. | A common phenotype referred to as “bulbar ALS” in previous phenotype classifications [7-13]. However, the term of “bulbar” does not describe a body region as such and has been replaced by “head region”. This term is linguistically consistent with arm, trunk and leg onset. |
| **distal arm onset** | O2d | – | Weakness or/and slowed, poorly coordinated voluntary movement in the hand muscles are first symptoms. | A common phenotype referred to as “limb ALS” in prior phenotype classifications [7-13]. However, previous classifications have not differentiated between distal vs proximal arm onset. It addresses research on the patterns of propagation following distal vs proximal arm onset. |
| **proximal arm onset** | O2p | – | Weakness in the shoulder muscles are first symptoms. | This phenotype is associated with an increased probability of developing the O2p, PL phenotype (syn.: flail-arm syndrome) [31, 43]. |
| **trunk respiratory onset** | O3r | diaphragmatic ALS, thoracic onset ALS | Weakness of respiratory muscles and hypoventilation presenting with orthopnea or dyspnea are first symptoms. | A rare phenotype which is difficult to classify clinically, as hypoventilation may be the only symptom. Previous classifications of trunk onset have rarely distinguished between respiratory and paravertebral onset [32]. |
| **trunk paravertebral onset** | O3p | dropped head syndrome; camptocormia | Weakness of paravertebral muscles of the neck and/or dorsal muscles are first symptoms. | A rare phenotype that is observed in clinical practice but rarely differentiated in previous phenotype classifications [32]. |
| **distal leg onset** | O4d | – | Weakness or/and slowed, poorly coordinated voluntary movement in the foot muscles are first symptoms. | A common phenotype referred to as “limb ALS” in prior phenotype classifications [7-13]. However, previous classifications did not distinguish between distal vs proximal leg onset. It addresses research on the patterns of propagation following distal vs proximal leg onset. |
| **proximal leg onset** | O4p | – | Weakness or/and slowed, poorly coordinated voluntary movement onset in the hip muscles are first symptoms. | See above. |
| **Propagation pattern** |  |  |  |  |
| **Earlier propagation** | PE | classic ALS | Propagation of slowed, poorly coordinated voluntary movements or weakness from the region of onset to another vertically distant body region *within 12 months* of symptom onset. | A common phenotype that was subsumed under “bulbar ALS” or “spinal ALS” in prior phenotype classifications [7-13]. |
| **Later propagation** | PL | It includes "progressive bulbar paralysis (PBP)", "flail arm-syndrome", and "flail leg-syndrome" (see below). | Propagation of slowed, poorly coordinated voluntary movements or weakness from the region of onset to another, vertically distant body region *later than 12 months* of symptom onset. | Rare phenotypes previously referred to as "progressive bulbar paralysis (PBP)", "flail arm-syndrome", and "flail leg-syndrome" [4, 9, 25, 43] (see below). |
|  |  | progressive bulbar or pseudobulbar paralysis | Refers to O1, PL | A rare phenotype previously referred to as "progressive bulbar paralysis (PBP)". Previous classifications have not consistently defined the time criterion to differentiate between "bulbar ALS" (O1, PE) and "PBP" (O1, PL). Several authors suggested the absence of limb involvement in the first 6 months as a criterion for P1I [9]. However, a median time of 12 months was found to distinguish "bulbar ALS" (O1, PE) from "PBP (O1, PL)" [25]. |
|  |  | flail arm syndrome, brachial amyotrophic diplegia; Vulpian-Bernhardt syndrome, hanging-arm syndrome, person-in-a-barrel syndrome, brachial amyotrophic diplegia [43] | Refers to O2d, PL and O2dp, PL | A rare phenotype previously referred to as "flail arm-syndrome". Most authors have suggested that this phenotype begins with proximal muscle weakness and atrophy [4]. However, distal onset and asymmetric presentations have been observed and can be classified as part of this phenotype. The main criterion to distinguish O2d/p, PE from O2d/p, PL is motor neuron dysfunction in the arms without relevant propagation to other regions within 12 months of symptom onset [4, 9, 43]. |
|  |  | flail leg syndrome, wasted leg syndrome, pseudopolyneuritic variant [30, 31]. | Refers to O4d, PL and O4dp, PL | A rare phenotype previously referred to as "flail leg-syndrome". Most authors have suggested that this phenotype begins with distal muscle weakness and atrophy. However, proximal onset has been observed and can be classified as part of this phenotype. The main criterion to distinguish O4d/p, PE from O4d/p, PL is motor neuron dysfunction in the legs without relevant propagation to other regions within 12 months of symptom onset [4, 9, 43]. |
| **Propagation not classifiable** | PN | – | Propagation of slowed, poorly coordinated voluntary movements or weakness from the region of onset to another, vertically distant body region not yet classifiable as time since symptom onset is less than 12 months. | An operational classification for a phenotype that is suggestive of PL with a disease duration of less than 12 months since symptom onset and therefore does not yet meet the time criterion to make the formal phenotype classification of PL [4, 9, 25, 43]. |

| **Motor neuron dysfunction** |  |  |  |  |
| --- | --- | --- | --- | --- |
| balanced motor neuron dysfunction | M0 | classic ALS [6-16] | A balanced combined MN dysfunction of the upper motor neuron (UMN) and the lower motor neuron (LMN) in any of the affected body regions is found. | A common phenotype referred to as "classic ALS" in previous phenotype classifications [6-16]. |
| dominant upper motor neuron (UMN) dysfunction | M1d | pyramidal ALS [4] | Dominant UMN dysfunction is found, presenting mainly with slowed, poorly coordinated voluntary movements, whereas discrete LMN dysfunction is also present. Each weakness is MRC grade 4 or greater [49]. | A rare phenotype previously referred to as "pyramidal ALS" [4]. This phenotype can be distinguished from M0 (classic ALS) by the extent of LMN signs. M0 shows weakness (MRC 4 or lower), whereas M1d is limited to discrete LMN dysfunction with low-grade atrophy and/or weakness (MRC 4 or -5) [4, 49]. |
| pure UMN dysfunction | M1p | primary lateral sclerosis, (PLS) [24, 25] | Pure UMN dysfunction is found, presenting mainly with slowed, poorly coordinated voluntary movements without apparent LMN dysfunction. The historic phenotype of PLS can diagnosed with certainty only after monitoring for LMN dysfunction for 48 months (M1p for 48 months) [6, 49]. | A rare phenotype previously referred to as primary lateral sclerosis, (PLS) [24, 25]. Bulbar symptoms, pseudobulbar affect, atrophy, weight loss, and any weakness are more predictive of M1d or M0 than M1p. Symmetry at presentation is possible in M1p and would be uncommon in M1d. The development of marked asymmetry over the course of the disease is expected [49]. |
| dominant lower motor neuron (LMN) dysfunction | M2d | – | Dominant LMN dysfunction is found, presenting mainly with weakness and associated atrophy, whereas discrete UMN dysfunction is also present. | A rare phenotype that can be distinguished from M1 by the extent of UMN signs. M3d is limited to preserved reflexes in atrophic and/or weak muscles. In contrast, hyperreflexia, and spread of reflexes to adjacent muscles are suggestive for M1. |
| pure LMN dysfunction | M2p | progressive muscle atrophy (PMA) [19-21] | Pure LMN dysfunction is found, presenting mainly with weakness and associated atrophy, without apparent UMN dysfunction. The historic phenotype of PMA can be diagnosed with certainty only after monitoring for UMN dysfunction for 48 months (M2p for 48 months) [4, 6]. | A rare phenotype previously referred to as progressive muscle atrophy (PMA) [19-21]. UMN dysfunction may develop in the later stages of the disease. Difficulties remain in correctly identifying the signs of UMN, especially in cases of mild involvement [16, 22, 23]. |
| dissociated motor neuron dysfunction | M3 | brachial amyotrophic spastic paraparesis variant of ALS [46, 47] | Dissociated MN dysfunction is found, presenting mainly with dominant LMN dysfunction of the arms and dominant UMN dysfunction of the legs. | A rare phenotype that is observed in clinical practice but not distinguished in previous phenotype classifications. |

### Assessment of temporal and spatial propagation

Phenotypes of propagation differentiate the temporal and spatial pattern in which motor neuron dysfunction spreads from the region of onset to another body region. The assessment is based on the patient history and/or neurological physical examinations in a follow-up of at least 12 months (**Figure 1 and 2**, **Table 1**).

### Assessment of motor neuron dysfunction

A neurological examination is performed to assess the symptoms and signs of UMN and/or LMN dysfunction. The clinical criteria for UMN and LMN dysfunction are related to the Gold Coast criteria for the diagnosis of ALS and are summarized in **Table 2** [44]. The evaluation is limited to a physical examination and excludes electrophysiologic, laboratory, and imaging studies. The assessment is based on a neurological examination follow-up (**Figure 1 and 2**, **Table 1**).

**Table 2.**
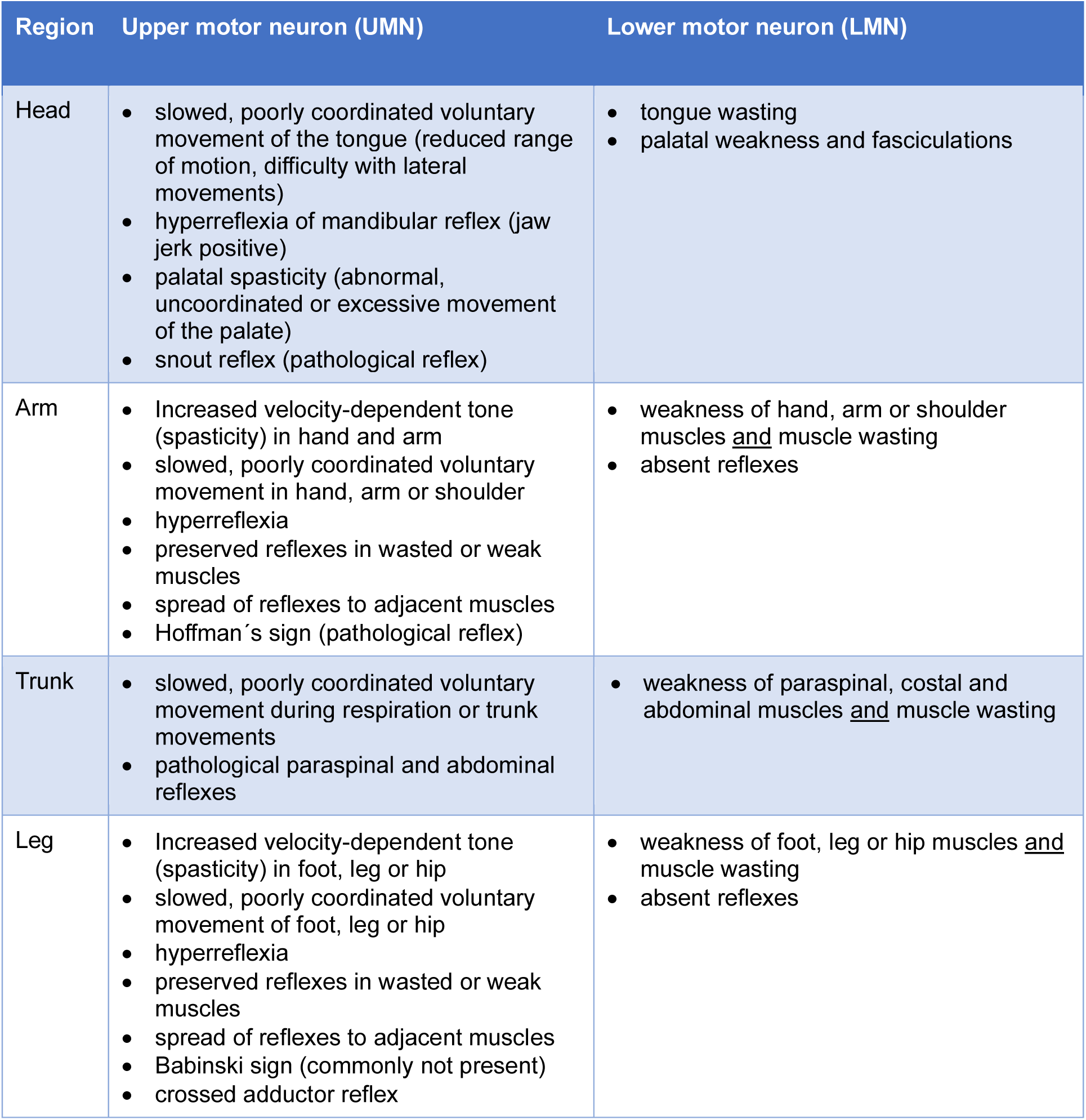
Motor neuron signs and symptoms. (according to Gold Coast criteria [41])

| Region | Upper motor neuron (UMN) | Lower motor neuron (LMN) |
| --- | --- | --- |
| Head | • slowed, poorly coordinated voluntary movement of the tongue (reduced range of motion, difficulty with lateral movements)<br>• hyperreflexia of mandibular reflex (jaw jerk positive)<br>• palatal spasticity (abnormal, uncoordinated or excessive movement of the palate)<br>• snout reflex (pathological reflex) | • tongue wasting<br>• palatal weakness and fasciculations |
| Arm | • Increased velocity-dependent tone (spasticity) in hand and arm<br>• slowed, poorly coordinated voluntary movement in hand, arm or shoulder<br>• hyperreflexia<br>• preserved reflexes in wasted or weak muscles<br>• spread of reflexes to adjacent muscles<br>• Hoffman's sign (pathological reflex) | • weakness of hand, arm or shoulder muscles <u>and</u> muscle wasting<br>• absent reflexes |
| Trunk | • slowed, poorly coordinated voluntary movement during respiration or trunk movements<br>• pathological paraspinal and abdominal reflexes | • weakness of paraspinal, costal and abdominal muscles <u>and</u> muscle wasting |
| Leg | • Increased velocity-dependent tone (spasticity) in foot, leg or hip<br>• slowed, poorly coordinated voluntary movement of foot, leg or hip<br>• hyperreflexia<br>• preserved reflexes in wasted or weak muscles<br>• spread of reflexes to adjacent muscles<br>• Babinski sign (commonly not present)<br>• crossed adductor reflex | • weakness of foot, leg or hip muscles <u>and</u> muscle wasting<br>• absent reflexes |

### Three-determinant classification of ALS motor phenotypes

Phenotypes were classified according to three anatomical determinants: 1) onset region (O); 2) temporal and spatial propagation of motor symptoms (P); and 3) the degree of UMN and/or LMN dysfunction (M) (**Table 1-2, Figure 2**). Accordingly, this ALS motor phenotype ordering system is referred to as the “OPM classification”. Examples of practical application of the OPM classification are provided in **Table 3**.

**Table 3.** ALS motor phenotype classification – examples of assessment.

| Example | Clinical description | Coded classification |
| --- | --- | --- |
| “Classic ALS, bulbar onset”, example #1 | History: onset with dysarthria → **O1**<br>Follow-up history: progressive dysarthria and, after 10 months from onset, weakness of right arm → **PE**<br>Investigation: weakness (LMN) and slowed, poorly coordinated voluntary movement (UMN) of the tongue and of right arm → **M0** | **O1, PE, M0** |
| “Classic ALS, bulbar onset”, example #2 | History: onset with dysarthria → **O1**<br>Follow-up history: progressive dysarthria and, after 14 months from onset, weakness of right arm → **PL**<br>Investigation: weakness of tongue and of right arm (LMN), discrete UMN symptoms → **M2d** | **O1, PL, M2d** |
| “Progressive bulbar paralysis (PBP)”, example #1 | History: onset with dysarthria → **O1**<br>Follow-up history: progressive dysarthria and, after 20 months from onset, no limb involvement → **PL**<br>Investigation: weakness and atrophy (LMN) of tongue; no UMN symptoms → **M2p** | **O1, PL, M2p** |
| “Progressive (pseudo)bulbar paralysis (PBP)”, example #2 | History: onset with dysarthria → **O1**<br>Follow-up history: progressive dysarthria and, after 8 months from onset, no limb involvement → **PN**<br>Investigation: slowed, poorly coordinated voluntary movement of the tongue; hyperreflexia of mandibular reflex; palatal spasticity (UMN); discrete wasting and fasciculations of the tongue (LMN) → **M2d** | **O1, PN, M1d** |
| “Classic ALS, arm onset”, example #1 | History: onset with weakness of left hand → **O2d**<br>Follow-up history: progressive weakness of left arm and, after 6 months from onset, dysarthria → **PE**<br>Investigation: weakness (LMN) and slowed, poorly coordinated voluntary movement (UMN) of the arm and weakness, atrophy of the tongue → **M0** | **O2d, PE, M0** |
| “Classic ALS, arm onset”, example #2 | History: onset with weakness of left hand → **O2d**<br>Follow-up history: progressive weakness of left arm and, after 22 months from onset, dysarthria → **PL**<br>Investigation: weakness (LMN) and slowed, poorly coordinated voluntary movement (UMN) of the arm and weakness, atrophy of the tongue → **M0** | **O2d, PL, M0** |
| “Classic ALS, arm onset”, example #3 | History: onset with weakness of left hand → **O2d**<br>Follow-up history: progressive weakness of left arm and, after 5 months from onset, weakness of left leg → **PE**<br>Investigation: weakness and atrophy (LMN) of the left arm and leg; preserved reflexes (discrete UMN) → **M2d** | **O2d, PE, M2d** |
| “Classic ALS, arm onset”, example #4 | History: onset with “stiffness” of left hand → **O2d**<br>Follow-up history: progressive slowed, poorly coordinated voluntary movement of left arm and, after 5 months from onset, “stiffness” of left leg → **PE**<br>Investigation: slowed, poorly coordinated voluntary movement and hyperreflexia (UMN) of the left arm and | **O2d, PE, M1d** |
|  | leg; discrete atrophy of the left hand and generalized fasciculations (discrete LMN) → **M1d** |  |
| “Flail-arm syndrome”, example #1 | History: onset with weakness of the proximal right arm → **O2p**<br>Follow-up history: progressive weakness of the right arm and, after 24 months from onset, weakness of the left shoulder → **PL**<br>Investigation: weakness and atrophy of bilateral shoulders (LMN), no UMN symptoms; no involvement of head, trunk and legs → **M2p** | **O2p, PL, M2p** |
| “Flail-arm syndrome”, example #2 | History: onset with weakness of the proximal right arm → **O2p**<br>Follow-up history: progressive weakness of the right arm and, after 10 months from onset, weakness of the left shoulder → **PN**<br>Investigation: weakness and atrophy of bilateral shoulders (LMN), no UMN symptoms; no involvement of head, trunk and legs → **M2p** | **O2p, PN, M2p** |
| “Respiratory ALS”, example #1 | History: onset with hypoventilation → **O3r**<br>Follow-up history: progressive hypoventilation and, after 7 months from onset, weakness of left arm → **PE**<br>Investigation: weakness and atrophy of the respiratory muscles and the arm (LMN); no UMN symptoms → **M2p** | **O3r, PE, M2p** |
| “Respiratory ALS”, example #2 | History: onset with hypoventilation → **O3r**<br>Follow-up history: progressive hypoventilation and, after 14 months from onset, weakness/instability of the trunk → **PL**<br>Investigation: weakness and atrophy of the respiratory muscles and the trunk (LMN); no UMN symptoms; no involvement of head, arms and legs → **M2p** | **O3r, PL, M2p** |
| “Dropped head syndrome” | History: onset with weakness/instability of the neck (dropped head) → **O3a**<br>Follow-up history: progressive weakness/instability of the neck and trunk and, after 6 months from onset, weakness of both arms → **PE**<br>Investigation: weakness and atrophy of axial muscles and bilateral shoulders (LMN); no UMN symptoms; no involvement of head and legs → **M2p** | **O3a, PE, M2p** |
| “Classic ALS, leg onset”, example #1 | History: onset with weakness of right foot → **O4d**<br>Follow-up history: progressive weakness of right leg and, after 8 months from onset, weakness of left arm → **PE**<br>Investigation: weakness (LMN) and slowed, poorly coordinated voluntary movement and hyperreflexia (UMN) of left arm and leg; weakness and atrophy of left leg → **M2p** | **O4d, PE, M2p** |
| “Classic ALS, leg onset”, example #2 | History: onset with weakness of left hip muscles → **O4p**<br>Follow-up history: progressive weakness of left leg and, after 13 months from onset, weakness of right leg and left arm → **PL** | **O4p, PL, M2d** |
|  | Investigation: weakness and atrophy (LMN) of bilateral legs and left arm; preserved reflexes in wasted muscle of the left hand → **M2d** |  |
| “Classic ALS, leg onset”, example #3 | History: onset with weakness of right foot → **O4d**<br>Follow-up history: progressive weakness of right leg and, after 8 months from onset, weakness of left leg → **PE**<br>Investigation: weakness (LMN) and slowed, poorly coordinated voluntary movement and hyperreflexia (UMN) of the arm; weakness and atrophy of left leg → **M0** | **O4d, PE, M2p** |
| “Flail-leg syndrome”, example #1 | History: onset with weakness of left foot → **O4p**<br>Follow-up history: progressive weakness of left leg and, after 20 months from onset, weakness of right leg → **PL**<br>Investigation: weakness and atrophy (LMN) of bilateral legs; no UMN symptoms → **M2p** | **O4d, PL, M2p** |
| “Flail-leg syndrome”, example #2 | History: onset with weakness of left foot → **O4d**<br>Follow-up history: progressive weakness of left leg and, after 11 months from onset, weakness of right leg → **PN**<br>Investigation: weakness and atrophy (LMN) of bilateral legs; preserved reflexes in wasted muscle of the left leg → **M2d** | **O4d, PN, M2d** |
| “pyramidal ALS”, example #1 | History: onset with “stiffness” of left hip and leg → **O4p**<br>Follow-up history: progressive slowed, poorly coordinated voluntary movement of left leg and, after 8 months from onset, “stiffness” of left arm and left leg → **PE**<br>Investigation: spasticity, and asymmetric, slowed, poorly coordinated voluntary movement and hyperreflexia (UMN) of the left arm and both legs; discrete atrophy of both hand muscles (discrete LMN) → **M1d** | **O4p, PE, M1d** |
| “pyramidal ALS”, example #2 | History: onset with dysarthria → **O1**<br>Follow-up history: progressive dysarthria and, after 10 months from onset, slowed, poorly coordinated voluntary movement of bilateral arms and legs → **PE**<br>Investigation: hyperreflexia of mandibular reflex; palatal spasticity, slowed, poorly coordinated voluntary movement of the tongue, arms and legs (UMN); discrete wasting and fasciculations of the tongue and hand muscles; asymmetric, low-grade weakness (MRC grade 4) of hip muscles (discrete LMN) → **M1d** | **O1, PE, M1d** |
| primary lateral sclerosis, (PLS), example #1 | History: onset with “stiffness” of both legs → **O4d**<br>Follow-up history: progressive slowed, poorly coordinated voluntary movement of both legs and, after 20 months from onset, asymmetric “stiffness” of both arms → **PL**<br>Follow-up investigation (after 48 months from onset): discrete dysarthria; hyperreflexia of mandibular reflex; palatal spasticity; high-grade spasticity, and asymmetric, highly slowed, poorly coordinated voluntary movement and hyperreflexia of all limbs; no LMN symptoms → **M1p** | **O4p, PL, M1p** |
| primary lateral sclerosis, (PLS), example #2 | History: onset with dysarthria → **O1**<br>Follow-up history: progressive dysarthria and, after 10 months from onset, slowed, poorly coordinated voluntary movement of bilateral arms and legs → **PE**<br>Follow-up investigation (after 48 months from onset): severe dysarthria; hyperreflexia of mandibular reflex; palatal spasticity; high-grade spasticity, and asymmetric, highly slowed, poorly coordinated voluntary movement and hyperreflexia of all limbs; no LMN symptoms → **M1p** | **O1, PE, M1p** |
| progressive muscle atrophy (PMA), example #1 | History: onset with weakness of right foot → **O4d**<br>Follow-up history: progressive weakness of left leg and, after 14 months from onset, asymmetric weakness of both legs and hands → **PL**<br>Follow-up investigation (after 48 months from onset): asymmetric weakness and wasting of foot, leg, hip, hand, arm and shoulder muscles; absent reflexes; no UMN symptoms → **M2p** | **O4d, PL, M2p** |
| progressive muscle atrophy (PMA), example #2 | History: onset with weakness of right left hand → **O2d**<br>Follow-up history: progressive weakness of left hand and, after 8 months from onset, asymmetric weakness of both arms, trunk, legs → **PE**<br>Follow-up investigation (after 14 months from onset): slight dysarthria, atrophy and weakness of the tongue; severe asymmetric weakness and wasting of limbs and trunk muscles; hypoventilation; absent reflexes; no UMN symptoms → **M2p** | **O2d, PE, M2p** |
| “brachial amyotrophic spastic paraparesis variant” of ALS | History: onset with weakness of right left hand → **O2d**<br>Follow-up history: progressive weakness of left hand and, after 14 months from onset, asymmetric weakness of both hands → **PL**<br>Follow-up investigation (after 20 months from onset): slight dysarthria, atrophy and weakness of the tongue; severe asymmetric weakness and wasting of hands and arms; no UMN symptoms of the upper limbs; high-grade spasticity, and asymmetric, highly slowed, poorly coordinated voluntary movement and hyperreflexia of both legs; no LMN symptoms of lower limbs → **M3** | **O2d, PL, M3** |

## DISCUSSION

This consensus process took the initiative to further evolve the classification of ALS motor phenotypes. Several common and rare ALS phenotypes are currently distinguished in everyday diagnosis and patient management. However, the description of phenotypes is informal, unsystematic and open to interpretation. Existing classifications were reported as inconsistent and insufficient to accurately describe ALS phenotypes [16, 17, 39]. Thus, phenotypes are mostly listed in the same rank order although they belong to different anatomical determinants. For example, “LMN dominant ALS”, “PMA” and “flail-arm syndrome” is often given as equal options for phenotype classification although the LMN dominant phenotype (M2d) and PMA (M2p) refer to the degree of motor neuron dysfunction, whereas flail-arm syndrome describes a different anatomical determinant – the propagation pattern (PL). From a research and trial perspective, it is imperative to clearly distinguish the anatomical axis of the phenotyping. As such, it is conceivable that investigational drugs will target selected phenotypic determinants (e.g. drugs acting on muscle cells, requiring inclusion of M2d, M2p and exclusion of M1d, M1p). Conversely, other phenotypes can be excluded to reduce clinical heterogeneity in clinical trials (e.g. exclusion of O3 and PL phenotypes). For both aspects, ensuring anatomical target engagement and controlling for heterogeneity, improved phenotype classification is needed.

### Three-determinant anatomical classification of phenotypes

This OPM classification was derived from an ongoing, large-scale multicenter biomarker study in which a two-determinant phenotype classification is being used [42]. The need for a revision of the current classification was driven by the practical experience of strengths and weaknesses inherent to the two-determinant classification. The OPM classification overcomes most of the constraints and incorporates several notable changes. These include the listing of agreed UMN/LMN symptoms, a more precise delineation of regions of onset, the addition of temporal criteria for the classification of propagation patterns, and the provision of a commented practical guideline for the classification of motor phenotypes (**Table 1 and 3**).

### Phenotypes of onset region

Previous classifications distinguished between bulbar, limb, and respiratory onset [4–17]. The term “bulbar” has been replaced with the term “head region”. This change was made for consistency, as the term “bulbar” does not describe a body region as such. “Head region” is linguistically consistent with arm, trunk, and leg onset. Another reason to abandon the term bulbar onset is the inconsistency that two different anatomical determinants, the region of onset and the (lower) motor neuron involvement, are intertwined in this term. On the same note, the wording of “bulbar onset” can be used for both, bulbar and pseudobulbar symptoms. In this revised classification, a more differentiated and, admittedly, “technical” terminology is introduced. Instead of the established term bulbar onset, it now reads “O1”, meaning onset in the head region (with dysarthria/dysphagia). The differentiation between balanced (M0), bulbar (M2) or pseudobulbar (M1) symptoms can be made in the phenotype determinant of motor neuron involvement. This distinction is of clinical relevance, as bulbar (M2) or pseudobulbar (M1) phenotypes have been associated with different prognoses and probabilities of concurrent frontotemporal dementia [28]. Furthermore, limb onset has been further differentiated, as it is conceivable that arm and leg onset, as well as distal (O2d, O4d) and proximal onset (O2p, O4p), may be associated with different disease courses and therefore of predictive value. In addition, trunk onset was subdivided into diaphragmatic and paravertebral onset – a differentiation that has not been realized in previous classifications. These rare variants may have different outcomes and need to be differentiated. Assessing the region of onset can be difficult. This information depends on the patient’s reflection, recollection, and interpretation. Also, the assessment is influenced by the neurologist’s interviewing skills to elicit the first symptoms. From the variety of early and prodromal signs and symptoms, the investigator is selecting the agreed onset symptom (dysarthria, dysphagia, limb paresis, or hypoventilation), and excluding symptoms that do not apply (fasciculations, cramps, and atrophy). Once the first symptom has been defined, the patient is asked for the onset date (year and month) of the agreed symptom. Given the retrospective nature of this assessment, there may be some level of uncertainty about the order of body regions involved, particularly in patients with faster progression and almost parallel motor neuron dysfunction in different regions. In general, phenotypes of onset regions have both, an investigator and a patient-reported component (**Figure 1**).

### Phenotypes of propagation pattern

Basically, two main dynamics of propagation are distinguished – earlier vs later propagation. Earlier propagation represents the typical course of disease in which motor dysfunction spreads from the site of onset to other regions within 12 months. In contrast, later propagation includes phenotypes in which motor symptoms are confined to the region of onset for at least 12 months. These temporal criteria for earlier vs later propagation is related to the diagnostic criteria of flail-arm (O2p, PL) and flail-leg (O4p, PL) syndromes which represent the major phenotypes for later propagation [45]. In both phenotypes, a follow-up period of at least 12 months has been defined to exclude the propagation of motor symptoms from the initially affected limb to other body regions. Also, in PBP (O1, PL), the same observation interval has been proposed [28]. Given the agreed temporal criteria for phenotyping later propagation (PL), these phenotypes cannot be assessed before 12 months after disease onset. In patients with symptoms confined to the region of onset and no clinical evidence of spread to other regions, the phenotype must be assessed as “not classifiable” (PN). This classification must also be applied to patients with unclear propagation pattern. In general, once the propagation phenotype has been assessed, the classification remains unchanged.

In the prior two-determinant classification, region of onset and propagation, were combined as one phenotype [42]. In the OPM classification, the propagation phenotype has been separated from the onset axis and has been introduced as an independent anatomical determinant (**Table1, Figure 2**). This distinction is based on the prognostic impact of both, the onset region, and the temporal propagation of motor neuron dysfunction. Thus, the region of onset alone does not predict the clinical course and prognosis. It is the temporal and spatial pattern in the spreading of motor dysfunction that is most relevant for the prognosis. For example, onset in the arm may result in rapid vertical spreading to the head, trunk or leg region (O2p/d, PE) or, conversely, to a horizontal spreading to the contralateral arm with protracted vertical spreading to other regions (O2p/d, PL; synonymous: flail-arm syndrome). Both scenarios start from the same region but are associated with significantly different trajectories. The predictive relevance of topography in terms of propagation pattern has also been described for head onset (O1, PE vs O1, PL; synonymous: bulbar onset ALS vs PBP) and leg onset (P4d/p, PE, P4p/d, PL; synonymous: classic ALS vs flail-leg syndrome) [28, 45]. The temporal criteria for distinguishing between early vs late vertical propagation in its historic descriptions are based on several observational studies [6, 11, 45]. However, the defined time frame of 12 months is rather arbitrary. Further studies will contribute to specify the point at which the trajectories of early and late propagation begin to bifurcate. Anatomical phenotypes were reported to have different contributions to the elevation in NfL that were independent of the different progression rates being associated with phenotypes. This observation contributed to the notion that phenotypes may be considered for future predictive models [42]. Thus, to reduce complexity in clinical trials, it may be conceivable to exclude the phenotypes with dominant horizontal spreading and prolonged vertical spread (O2p/d, PL, flail-arm syndrome; O4p/d, PL, flail-leg syndrome), which are known to have significantly different trajectories, including longer survival [8–15, 42].

### Phenotypes of motor neuron dysfunction

Previously reported phenotypes of motor neuron dysfunction included balanced, dominant, and pure UMN and/or LMN involvement [42]. In this revised classification, the phenotype of dissociated motor neuron dysfunction (M3) has added. The M3 phenotype presents with dominant or pure LMN dysfunction of the arms and dominant or pure UMN dysfunction of the legs – previously referred to as the brachial amyotrophic spastic paraparesis variant of ALS [46, 47]. In clinical practice, this phenotype is mostly found in patients with early adult onset being associated with a prolonged disease course. However, systematic data on the frequency, the correlation with NfL levels, and the prognosis of this phenotypic variant are lacking and will be the subject of future investigations.

The historic terms of PMA and PLS are widely used and established. However, in this classification, a more systematic ordering was introduced in which PMA and PLS were only listed as secondary terms. The assessment of the motor neuron involvement phenotype is based on the neurological examination. As such, the classification of UMN and/or LMN dysfunction may be subject to considerable interobserver variability. For example, a reflex that is objectively reduced in the context of severe muscle atrophy might be regarded as brisk by an experienced neurologist [16]. Thus, the determinant of motor neuron involvement may be influenced by the level of clinical experience of the phenotype rater. The greatest inter-rater variability is expected in the distinction between balanced (M0) vs UMN dominant (M1d) and LMN dominant (M2d), respectively. As a limitation, no clinical grading of UMN and LMN symptoms exists. However, the implementation of this revised phenotype classification in controlled studies will determine the differences in grading between experienced and less experienced raters and the extent to which interrater variability of motor neuron involvement appears to be a challenge.

As the disease progresses, motor neuron symptoms change. Therefore, follow-up examinations are expected to reveal evolving degrees of UMN and LMN involvement, leading to a different phenotype classification (**Figure 1**). In advanced disease stages, LMN symptoms may dominate and mask UMN symptoms. Thus, a previously balanced phenotype (M0) may change to a dominant (M2d) or even pure (M2p) LMN involvement phenotype. Conversely, patients with pure LMN (M2p) or UMN (M1p) may later develop combined motor neuron symptoms resulting in the phenotype of M2d or M1d, respectively.

Monitoring for combined symptoms has been required before the phenotype of PLS or PMA can be classified in its historical description. Thus, for the diagnosis of PLS a monitoring of LMN symptoms between 3 and 5 years has been proposed, while Singer and Gordońs recommendation of monitoring for 48 months has been incorporated into the guidelines of this classification (**Table 1**) [48, 49]. Similarly, in the phenotyping of PMA, a monitoring period of 48 months (for UMN symptoms) has been defined [6]. These temporal criteria result in a more homogenous definition of PLS and PMA but at the same time, these phenotypes are tilted towards a longer course, less progression and higher survival. In a more open approach, the OPM classification allows to classify pure UMN/LMN independent of the temporal PLS/PMA criteria of 48 months. Therefore, this revised classification will be more inclusive for M1p and M2p patients of higher progression and shorter disease duration. In general, few studies have focused on the proportions of patients with dominant or pure UMN/LMN symptoms and the dynamics of phenotypes in terms of evolving degrees of motor neuron dysfunction. Implementation of the revised classification in trials or clinical practice will generate longitudinal data on changing motor phenotypes. These data may be relevant in the context of trials of investigational drugs that more specifically target upper or lower motor neurons.

### Challenges and further research in motor phenotyping

This OPM classification addresses a long-standing need for harmonization of motor phenotypes in ALS. The advantage of this classification lies in the generic nature of this ordering system, which also allows flexible modification and, if necessary, further differentiation of additional phenotypes. The authors acknowledge that the OPM phenotyping is constrained to motor phenotypes, with all non-motor phenotypes remaining unconsidered. The restriction to motor phenotypes has been instrumental in mitigating the complexity inherent in the development of a novel phenotypic classification. The main challenge is the implementation of the OPM phenotypes in clinical practice and trials. Thus, acceptance of the new, more technical terminology (e.g. M2p for PMA, M1p for PLS) among neurologists remains uncertain. This general concern and potential limitation also apply to other historical phenotypic variants of ALS, known as flail-arm (O2p/d, PL) and flail-leg (O4p/d, PL) syndromes. In general, historic names were preserved in respect to the achievement of prior generations and to support clinicians to aligning the new, more technical terms, with the historical descriptive wording. A further rationale for integrating the OPM classification with the historical terms pertains to the retrospective classification of phenotypes from existing data, wherein solely the historical terms were employed. The semantic alignment is the basis for manual retrospective classification, and even more importantly, in leveraging AI-based large language models to undertake this task. The historic terms (PMA, PLS, flail-arm and flail-leg syndrome) are expected to be continuously used in clinical practice. Realistically, a longer-term consensus and adjustment process is required to integrate the OPM phenotype classification in clinical practice. In the more formalized setting of clinical trials, a faster integration of the revised phenotypes is realistic. A first application of the revised classification is planned in an amendment of an ongoing multi-center study in which the OPM classification will be associated with ALS progression, serum NfL and survival [42]. This phenotype classification is limited to the symptomatic period of ALS where weakness is the defining criterion [50]. The definition of onset as weakness and/or slow and uncoordinated movements was linked to the ALS progression rate, also calculated with reference to the first motor symptoms or deficits [42, 43]. In further iterations of ALS phenotype classifications, the inclusion of the presymptomatic period (atrophy, fasciculations, spasms), laboratory findings (EMG, MRI) and biomarker signals (prodromal elevation of NfL) must be considered. Furthermore, the restriction to motor symptoms and the omission of the non-motor dimension of the disease spectrum must be understood from an operational point of view to reduce the complexity in the implementation of this classification. For this OPM classification alone, a step-by-step process of adaptation and improvement is expected. In further steps of the ALS phenotype classification, the integration of behavioral and cognitive phenotypes must be achieved.

## CONCLUSIONS

The OPM classification corresponds to a long-term objective of structuring the variable ALS motor phenotypes, with a value proposition on three different levels. First, in clinical practice, the phenotypes should contribute to further individualize the prognosis in the expected course, complementing other established prognostic factors such as the ALS progression rate, NfL, body mass index, or respiratory parameters. Second, in clinical trials, phenotypes will be helpful to decipher the different trajectories of ALS. This distinction may be increasingly important to reduce clinical heterogeneity of trial populations or, conversely, to target drug trials to specific phenotypes. Consequently, specific phenotypes in the OPM classification may be of interest for predictive models used for inclusion, exclusion, or stratification criteria in clinical trials. Third, focality and spreading are central features of ALS and important for the precise understanding of the disease whereas two basic mechanisms of spreading are discussed: contiguous and distant axonal spreading [1–3, 36]. The differentiation of phenotypes may also facilitate the link between clinical presentation, pathological anatomy and underlying molecular spreading mechanisms.

## Data Availability

All data produced in the present work are contained in the manuscript

## List of abbreviations

ALS: amyotrophic lateral sclerosis
FAS: flail-arm syndrome
FLS: flail-leg syndrome
LMN: lower motor neuron
OPM: onset, propagation, motor neuron dysfunction
PBP: progressive bulbar palsy
PLS: primary lateral sclerosis
PMA: progressive muscle atrophy
UMN: upper motor neuron

## DECLARATIONS

### Ethics approval and consent to participate

Not applicable

### Consent for publication

All authors of the paper agree with the manuscript and meet the criteria for authorship.

### Availability of data and materials

Not applicable

### Authors’ contributions

TM, MB, JG, AL, AM and TG made substantial contributions to the conception of the work; TM, MB, JG, PW, SB, PR, RS, AR, JW, UW, AL, JW, SP, PL, RG, WL, MW, CM, AM and TG contributed to the interpretation of existing data; TM, MB, JG, AL, PW, PR, AR, SP, PL, RG, WL, MW, AM and TG have drafted the work or substantively revised it.

## Acknowledgements

The authors wish to thank the Boris Canessa ALS Stiftung (Düsseldorf, Germany) for funding this work and continuous support.

## OPM classification of ALS motor phenotypes – commentary for the use in clinical practice and research

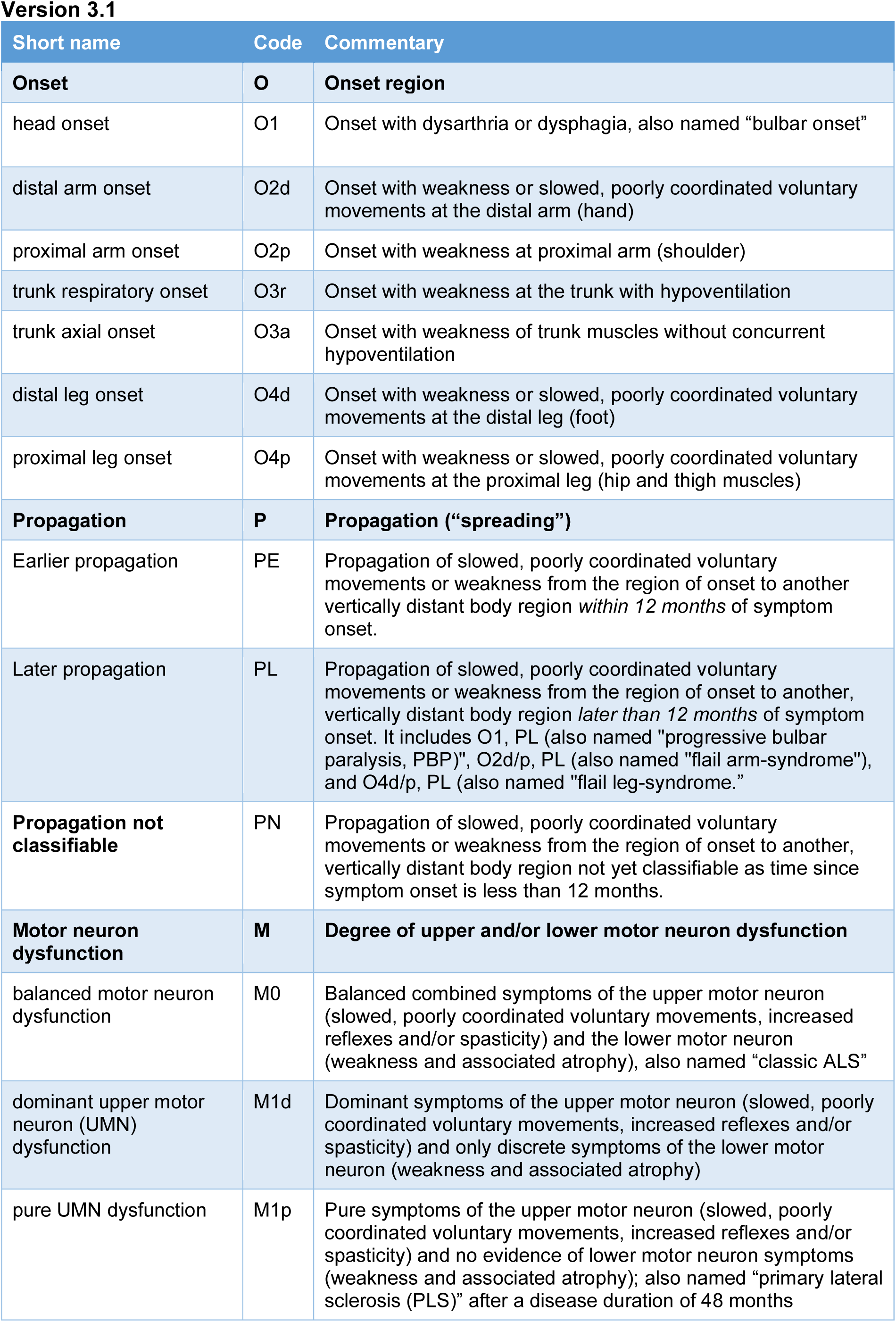
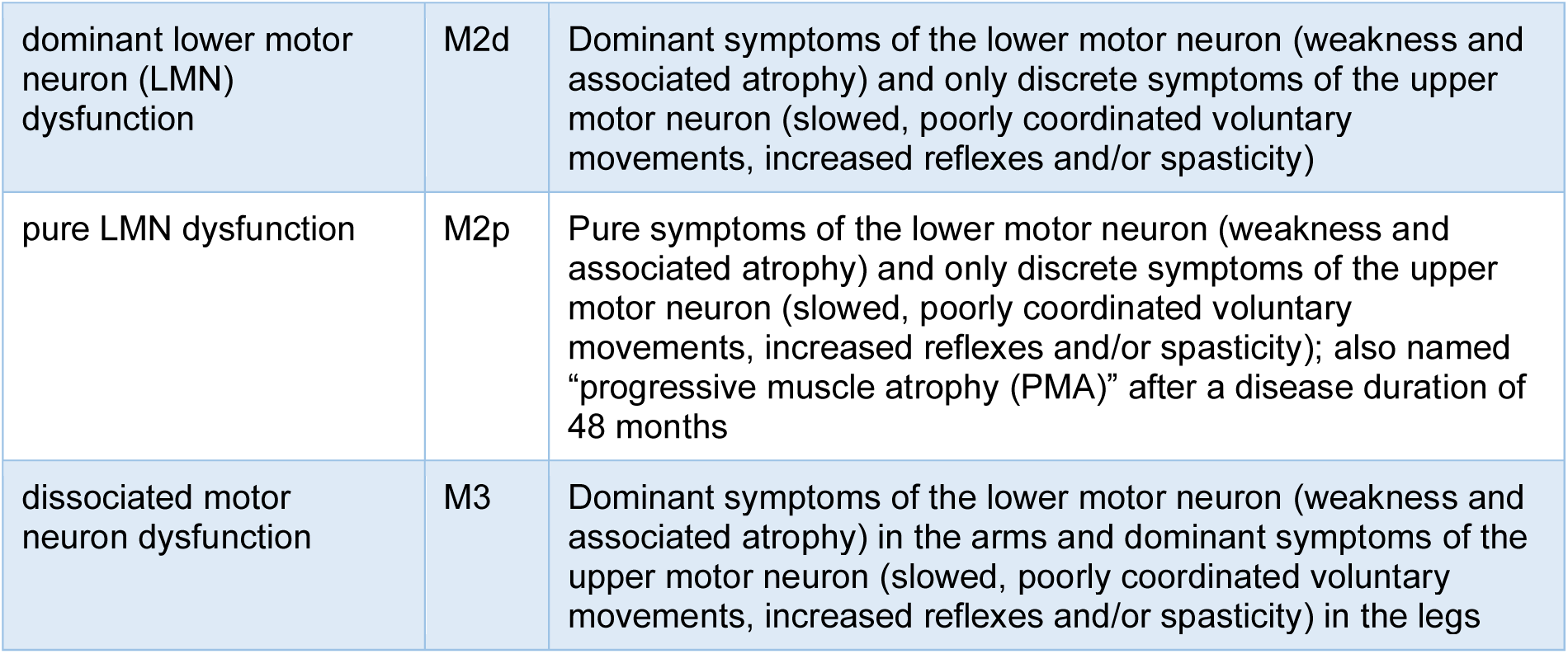

Version 3.1
| Short name | Code | Commentary |
| --- | --- | --- |
| **Onset** | **O** | **Onset region** |
| head onset | O1 | Onset with dysarthria or dysphagia, also named “bulbar onset” |
| distal arm onset | O2d | Onset with weakness or slowed, poorly coordinated voluntary movements at the distal arm (hand) |
| proximal arm onset | O2p | Onset with weakness at proximal arm (shoulder) |
| trunk respiratory onset | O3r | Onset with weakness at the trunk with hypoventilation |
| trunk axial onset | O3a | Onset with weakness of trunk muscles without concurrent hypoventilation |
| distal leg onset | O4d | Onset with weakness or slowed, poorly coordinated voluntary movements at the distal leg (foot) |
| proximal leg onset | O4p | Onset with weakness or slowed, poorly coordinated voluntary movements at the proximal leg (hip and thigh muscles) |
| **Propagation** | **P** | **Propagation (“spreading”)** |
| Earlier propagation | PE | Propagation of slowed, poorly coordinated voluntary movements or weakness from the region of onset to another vertically distant body region *within 12 months* of symptom onset. |
| Later propagation | PL | Propagation of slowed, poorly coordinated voluntary movements or weakness from the region of onset to another, vertically distant body region *later than 12 months* of symptom onset. It includes O1, PL (also named “progressive bulbar paralysis, PBP”), O2d/p, PL (also named “flail arm-syndrome”), and O4d/p, PL (also named “flail leg-syndrome.”) |
| **Propagation not classifiable** | PN | Propagation of slowed, poorly coordinated voluntary movements or weakness from the region of onset to another, vertically distant body region not yet classifiable as time since symptom onset is less than 12 months. |
| **Motor neuron dysfunction** | **M** | **Degree of upper and/or lower motor neuron dysfunction** |
| balanced motor neuron dysfunction | M0 | Balanced combined symptoms of the upper motor neuron (slowed, poorly coordinated voluntary movements, increased reflexes and/or spasticity) and the lower motor neuron (weakness and associated atrophy), also named “classic ALS” |
| dominant upper motor neuron (UMN) dysfunction | M1d | Dominant symptoms of the upper motor neuron (slowed, poorly coordinated voluntary movements, increased reflexes and/or spasticity) and only discrete symptoms of the lower motor neuron (weakness and associated atrophy) |
| pure UMN dysfunction | M1p | Pure symptoms of the upper motor neuron (slowed, poorly coordinated voluntary movements, increased reflexes and/or spasticity) and no evidence of lower motor neuron symptoms (weakness and associated atrophy); also named “primary lateral sclerosis (PLS)” after a disease duration of 48 months |
| dominant lower motor neuron (LMN) dysfunction | M2d | Dominant symptoms of the lower motor neuron (weakness and associated atrophy) and only discrete symptoms of the upper motor neuron (slowed, poorly coordinated voluntary movements, increased reflexes and/or spasticity) |
| pure LMN dysfunction | M2p | Pure symptoms of the lower motor neuron (weakness and associated atrophy) and only discrete symptoms of the upper motor neuron (slowed, poorly coordinated voluntary movements, increased reflexes and/or spasticity); also named "progressive muscle atrophy (PMA)" after a disease duration of 48 months |
| dissociated motor neuron dysfunction | M3 | Dominant symptoms of the lower motor neuron (weakness and associated atrophy) in the arms and dominant symptoms of the upper motor neuron (slowed, poorly coordinated voluntary movements, increased reflexes and/or spasticity) in the legs |

